# Leveraging Machine Learning Approaches to Identify Health-Related Social Needs Screening from Electronic Health Records

**DOI:** 10.64898/2026.06.23.26356305

**Authors:** Levente Dojcsak, Tadesse Abegaz, Mohaimenul Islam, Young Chandler, Arati Maleku, Anna Doubeni, Buhari Mohammed, Michael A. Langston, Macarius Donneyong

## Abstract

Health-related social needs (HRSNs), such as housing instability, food insecurity, and transportation challenges, are nonmedical factors associated with poorer health and well-being. Screening for unmet HRSNs is a critical step towards identifying at-risk patients, but manual screening is resource intensive and often incomplete. We utilized Electronic Health Records (EHR) data to develop machine learning models to identify unmet HRSNs using a limited set of non-modifiable sociodemographic features available in EHRs. We included 745,975 patients screened for at least one HRSN using data from community health centers that participated in the OCHIN practice-based research network between 2016 and 2022. Logistic regression, random forest (RF), eXtreme Gradient Boosting (XGBoost), and Light Gradient Boosting Machine (LightGBM) algorithms were trained to predict unmet HRSNs. Model performance was evaluated using 10-fold cross-validation and area under the receiver operating characteristic curve (AUROC). For overall HRSN prediction, LightGBM (AUROC, 64.5%, 95%CI: 64.3, 64.7) performed slightly better than logistic regression (61.4%), RF (63.7%), and XGBoost (60.3%). Similar performances were observed predicting individual HRSNs. Model performances were modest; however, they establish a benchmark for predictive performance achievable using only routinely available demographic data and provide a foundation for incorporating additional clinical and area-level social determinants of health data.

## INTRODUCTION

Unmet health-related social needs (HRSNs) are the manifestations of the societal and environmental conditions in which individuals are born, grow, live, work, and age (contextual social determinants of health [SDOH]) [1, 2]. These non-medical factors are associated with poorer health and well-being, and as such are targets of intervention in order to improve population health outcomes [3–5]. Although accurate and routine screening is a critical step towards addressing HRSNs, most healthcare providers tend to lack the capacity and resources to achieve this when caring for large patient populations.

Community health centers (CHCs) serve as safety-net clinics and provide vital and critical care to socioeconomically vulnerable and underinsured patients who would otherwise not be able to access care from traditional providers. The prevalence of unmet HRSNs is highest among the CHC populations. The Health Resources and Services Administration (HRSA), which funds CHCs, recommends that CHCs screen and address HRSNs [6]. CHCs, however, lack the capacity and resources to meet this mandate [7].

CHCs and other providers currently screen for HRSNs manually using questionnaires such as the Accountable Health Communities’ Health-Related Social Needs (AHC-HRSN) Tool, the Protocol for Responding to and Assessing Patients’ Assets, Risks, and Experiences (PRAPARE), the Health Leads Screening Toolkit, and the Centers for Medicare & Medicaid Services (CMS) Innovation Center’s Social Needs Screening Tools. However, the lack of standardization among these questionnaires creates inconsistencies in how unmet HRSNs are screened and documented across different healthcare settings, resulting in missed opportunities for early intervention [8–10]. Also, many of these questionnaires do not capture all the domains of HRSNs [11]. For example, some CHCs may focus screening on a subset of HRSNs that they are better equipped to address, thus contributing to missed identification of patients with unmet needs outside of those domains. Further, integrating these manual screening processes into clinical workflows is resource intensive and thus not scalable, especially for low-resourced CHCs [12]. While standardized screenings are ideal, we recognize that systematic screenings are difficult to implement.

To address the aforementioned challenges, we sought to explore the application of Machine Learning (ML) techniques to help identify patients with unmet HRSNs from Electronic Health Records (EHR) data. The challenges outlined above are not a comprehensive list of potential barriers to address HRSNs. While ML models cannot directly overcome systemic barriers such as limited resources or care coordination challenges, they can enhance the efficiency and accuracy of the screening process. ML-based HRSN prediction models, if integrated into EHR platforms, could potentially facilitate a seamless incorporation of routine HRSN screening into clinical workflows by identifying at risk patients, thus enabling healthcare organizations to allocate resources more effectively and lead to improved connectivity of patients to social intervention services and resources.

## METHODS

### Data Source

The data for this study was sourced from the OCHIN network [13, 14], a national network of community-based healthcare organizations that began in Oregon and later expanded to other states. OCHIN hosts and manages a centralized Epic EHR database system for over 300 healthcare organizations within its network. This EHR database captures a wide range of patient- and encounter-level data including demographics, encounter details, vital signs, diagnoses, procedures, immunizations, laboratory tests, medications, and other patient-reported outcomes. OCHIN’s extensive network currently includes over 2100 clinics across 41 states. In addition, the data from the Accelerating Data Value Across a National Community Health Center Network (ADVANCE) Clinical Research Network (part of PCORnet [15]) provided a comprehensive and diverse dataset for our study.

### Ethical Considerations

This study was approved by The Ohio State University Institutional Review Board (IRB). The requirement for patient informed consent was waived by The Ohio State University IRB because the data were de-identified by OCHIN and posed minimal risk of re-identification. This study was reported in accordance with the Transparent Reporting of a Multivariable Prediction Model for Individual Prognosis or Diagnosis (TRIPOD) statement [16]. All methods were performed in accordance with the relevant guidelines and regulations.

### Current HRSN Screening Strategies

The CHCs within OCHIN screen for several domains of unmet HRSNs (housing instability, inadequate housing quality, relationship safety, social isolation, transportation challenges, utility needs, food insecurity) using a variety of different questionnaires that in total involve more than120 question items. Screening is conducted according to each clinic’s individual protocols, and questionnaires may be administered to patients during routine visits or other encounters. For example, some clinics’ screening process for food insecurity involves the use of validated, evidence-based questions such as whether patients have “worried about running out of food” or “experienced food running out before they were able to afford more within the past 12 months” [17, 18]. For housing instability, patients may be asked whether they currently have stable housing, e.g., “What is your housing situation today?” Questions about poor housing quality, on the other hand, assess the safety and adequacy of their living conditions, e.g., “Does your housing have issues such as mold, pests, lead, or inadequate heating or cooling?” For inability to pay utility bills, patients may be asked whether they have experienced challenges in paying for utility services (such as electricity, heating, or water) within the past 12 months [9, 19]. Social isolation questions may inquire about social connections, frequency of interactions, feelings of loneliness, and the absence of supportive social relationships [20, 21]. HRSN screening responses from these various questionnaires were documented by participating clinics within the EHR and stored in structured categories, which enabled us to create datasets for use in this study. Beyond screening for HRSNs, some clinics may also ask patients directly whether they want the clinic’s help with addressing their social needs. For more details on how HRSN domains were selected and included in the database managed by OCHIN, please refer to Gold et. al. (2017) [22].

### Patient Selection

This study included all individuals who were screened for at least one HRSN between January 1, 2016, and December 31, 2022, and documented in the OCHIN EHR database. Although some patients underwent multiple screenings during this period, only data from their most recent HRSN screening were included in analysis.

### Outcome Variables

Unmet HRSNs, overall, and by domains, were the primary outcome variables for the ML prediction. A domain-specific binary outcome variable for unmet HRSN (present vs absent) was defined for each patient based on the presence of a positive response to at least one question item per domain. The overall HRSN binary outcome was defined as the composite of all the seven domains of HRSNs, i.e., the presence of at least one unmet HRSN. We focused on the following seven domains because they are among the most frequently reported unmet HRSNs [23]: food insecurity, housing instability, utility needs, transportation barriers, social isolation, relationship safety, and poor housing quality.

### Potential Predictors

We included the following non-modifiable person-level sociodemographic factors as potential predictors of unmet HRSNs: age, sex, race, ethnicity, marital status, sexual orientation, preferred language, and migration/seasonal status. These features were selected because they are readily available in most EHR databases, although not all are consistently captured across EHR systems, and they have been previously identified as potential risk factors of unmet HRSNs [24–26]. Prior work has shown that these sociodemographic factors are associated with disparities in access to care and social needs, influencing the likelihood of experiencing unmet HRSNs. Age, sex, marital status, race/ethnicity, and language have all been linked to differences in social risk exposure and health outcomes across populations [24–26]. We did not include other common EHR-based features such as medical diagnoses, healthcare utilization, and medication use because these factors reflect access to healthcare and are influenced by HRSNs, not the other way around. Although including them could improve model predictability, it would likely reduce interpretability by conflating social and clinical predictors. Moreover, at least one prior study has shown that adding such variables does not necessarily improve overall model accuracy [27]. We also excluded modifiable socioeconomic administrative variables such as federal poverty level (FPL) and insurance status, as our modeling approach was restricted to non-modifiable patient characteristics. Our goal was to develop parsimonious models that achieved comparable performance with fewer features.

### Statistical Analysis

#### Overview

The modeling approach involved three main tasks: data preprocessing, ML model development, and interpretation of the ML models. We reported the distribution of all the input features by each HRSN domain prior to implementing the ML analysis. All analyses were performed using Python (version 3.12).

#### Data Preprocessing

Data preprocessing involved (1) quality control and missing data assessment, (2) transformation of potential predictors, and (3) the construction of datasets for predicting the overall and domain-specific HRSN.

#### Quality control and missing data assessment

Missing values were defined as either unrecorded information or entries documented as “unknown” (**Table 1**). The proportion of missing data varied across features, ranging from 0.1% for sex to 84.4% for migration status. For variables with very low missingness (e.g. age, sex), samples with missing values were removed. Imputation was not performed, as the dataset was sufficiently large. For features with higher missingness, such as migration/seasonal status, we categorized missing values as “unknown.”

**Table 1.** Distribution of sociodemographic and healthcare-related characteristics of OCHIN network patients by overall and domain-specific health-related social needs (HRSNs) screened between January 1, 2016, and December 31, 2022. This cross-sectional predictive analysis included all 745,975 patients with at least one HRSN screening. For patients with multiple screenings, only the most recent was included. Overall HRSN status was defined as having at least one unmet HRSN in any of the seven domains: food insecurity, housing instability, poor housing quality, relationship safety, social isolation, transportation barriers, and utility needs. Because not all patients were screened for every domain, domain-specific columns are based on the subset of patients screened for that domain (N). Demographic characteristics are reported only for patients who screened positive for the corresponding unmet need (n). The total patients screened column summarizes the full cohort. For marital status, the “others” category includes domestic partner, significant other, widowed, and responses not otherwise classified. For sexual orientation, the “others” category includes asexual, queer, multiple orientations, something else, declined to answer, and unknown. Migration status indicates whether a patient was identified as a seasonal worker. The “unknown” category includes cases for which this information was not documented in the EHR. Values are listed as counts and percentages of the total, except age which is presented as mean and standard deviation (SD).

|  | Food Insecurity<br>(n=127,611;<br>screened<br>N=536,915) | Housing Instability<br>(n=85,555;<br>screened<br>N=464,156) | Housing Quality<br>(n=23,949;<br>screened<br>N=303,403) | Relationship Safety<br>(n=25,872;<br>screened<br>N=397,093) | Social Isolation<br>(n=38,126;<br>screened<br>N=220,455) | Transportation<br>(n=72,496;<br>screened<br>N=448,976) | Utility<br>(n=75,335;<br>screened<br>N=382,146) | Overall HRSN status<br>(n=199,920;<br>screened<br>N=745,975) | Total patients screened<br>(N=745,975) |
| --- | --- | --- | --- | --- | --- | --- | --- | --- | --- |
| **Age (Mean, SD)** | 38.7 (18.2) | 40.6 (16.6) | 39.7 (18.5) | 37.9 (15.5) | 41.5 (15.8) | 41.4 (17.1) | 40.6 (17.2) | 39.0 (18.3) | 37.4 (19.4) |
| **Sex** |  |  |  |  |  |  |  |  |  |
| Female | 70056 (54.9) | 44188 (51.6) | 14108 (58.9) | 17380 (67.2) | 21607 (56.7) | 38460 (53.1) | 42526 (56.4) | 113214 (56.6) | 467183 (62.6) |
| Male | 57370 (45.0) | 41286 (48.3) | 9803 (40.9) | 8459 (32.7) | 16479 (43.2) | 33970 (46.9) | 32747 (43.5) | 86457 (43.2) | 278267 (37.3) |
| Others | 185 (0.1) | 81 (0.1) | 38 (0.2) | 33 (0.1) | 40 (0.1) | 66 (0.1) | 62 (0.1) | 249 (0.1) | 525 (0.1) |
| **Marital status** |  |  |  |  |  |  |  |  |  |
| Single | 44067 (34.5) | 31714 (37.1) | 8598 (35.9) | 9516 (36.8) | 14570 (38.2) | 27821 (38.4) | 26696 (35.4) | 69217 (34.6) | 228172 (30.6) |
| Divorced/<br>Separated | 8090 (6.3) | 6142 (7.2) | 1785 (7.5) | 1969 (7.6) | 3122 (8.2) | 5584 (7.7) | 5448 (7.2) | 12767 (6.4) | 34023 (4.6) |
| Married | 13136 (10.3) | 8503 (9.9) | 2825 (11.8) | 2524 (9.8) | 4687 (12.3) | 6755 (9.3) | 8699 (11.5) | 22359 (11.2) | 117534 (15.8) |
| Others | 62318 (48.8) | 39196 (45.8) | 10741 (44.8) | 11863 (45.9) | 15747 (41.3) | 32336 (44.6) | 34492 (45.8) | 95577 (47.8) | 366246 (49.1) |
| **Race** |  |  |  |  |  |  |  |  |  |
| Asian | 3025 (2.4) | 1547 (1.8) | 342 (1.4) | 347 (1.3) | 621 (1.6) | 1315 (1.8) | 1469 (1.9) | 5020 (2.5) | 30810 (4.1) |
| Black | 36601 (28.7) | 25308 (29.6) | 6105 (25.5) | 5709 (22.1) | 8457 (22.2) | 21500 (29.7) | 22984 (30.5) | 55680 (27.9) | 172281 (23.1) |
| White | 69244 (54.3) | 47201 (55.2) | 14313 (59.8) | 16336 (63.1) | 24361 (63.9) | 40508 (55.9) | 41590 (55.2) | 110418 (55.2) | 427099 (57.3) |
| Others | 7137 (5.6) | 4487 (5.2) | 1461 (6.1) | 1494 (5.8) | 1921 (5.0) | 3668 (5.1) | 3675 (4.9) | 10988 (5.5) | 43743 (5.9) |
| Unknown | 11604 (9.1) | 7012 (8.2) | 1728 (7.2) | 1986 (7.7) | 2766 (7.3) | 5505 (7.6) | 5617 (7.5) | 17814 (8.9) | 72042 (9.7) |
| **Ethnicity** |  |  |  |  |  |  |  |  |  |
| Non-Hispanic | 85307 (66.8) | 60780 (71.0) | 17534 (73.2) | 20088 (77.6) | 29057 (76.2) | 53489 (73.8) | 54458 (72.3) | 136509 (68.3) | 460664 (61.8) |
| Hispanic | 35585 (27.9) | 20042 (23.4) | 5105 (21.3) | 4036 (15.6) | 6852 (18.0) | 14987 (20.7) | 17099 (22.7) | 52272 (26.1) | 240249 (32.2) |
| Others | 6719 (5.3) | 4733 (5.5) | 1310 (5.5) | 1748 (6.8) | 2217 (5.8) | 4020 (5.5) | 3778 (5.0) | 11139 (5.6) | 45062 (6.0) |
| **Sexual orientation** |  |  |  |  |  |  |  |  |  |
| Straight | 83881 (65.7) | 60151 (70.3) | 16359 (68.3) | 18373 (71.0) | 29112 (76.4) | 51530 (71.1) | 54353 (72.1) | 134832 (67.4) | 508808 (68.2) |
| Bisexual | 3811 (3.0) | 2566 (3.0) | 852 (3.6) | 1372 (5.3) | 1520 (4.0) | 2297 (3.2) | 2130 (2.8) | 5982 (3.0) | 15440 (2.1) |
| Gay/Lesbian | 1189 (0.9) | 807 (0.9) | 245 (1.0) | 352 (1.4) | 438 (1.1) | 722 (1.0) | 723 (1.0) | 1860 (0.9) | 5646 (0.8) |
| Others | 38730 (30.4) | 22031 (25.8) | 6493 (27.1) | 5775 (22.3) | 7056 (18.5) | 17947 (24.8) | 18129 (24.1) | 57246 (28.6) | 216081 (29.0) |
| **Migration status** |  |  |  |  |  |  |  |  |  |
| Yes | 224 (0.2) | 99 (0.1) | 23 (0.1) | 23 (0.1) | 28 (0.1) | 133 (0.2) | 95 (0.1) | 352 (0.2) | 2821 (0.4) |
| No | 20679 (16.2) | 13195 (15.4) | 3617 (15.1) | 3653 (14.1) | 5801 (15.2) | 10743 (14.8) | 11570 (15.4) | 30908 (15.5) | 133695 (17.9) |
| Unknown | 106708 (83.6) | 72261 (84.5) | 20309 (84.8) | 22196 (85.8) | 32297 (84.7) | 61620 (85.0) | 63670 (84.5) | 168660 (84.4) | 609459 (81.7) |
| **Language** |  |  |  |  |  |  |  |  |  |
| English | 95925 (75.2) | 68109 (79.6) | 20159 (84.2) | 23721 (91.7) | 33382 (87.6) | 59756 (82.4) | 60872 (80.8) | 154433 (77.2) | 537372 (72.0) |
| Spanish | 25501 (20.0) | 13594 (15.9) | 3026 (12.6) | 1665 (6.4) | 3791 (9.9) | 9815 (13.5) | 11386 (15.1) | 35562 (17.8) | 160069 (21.5) |
| Others | 6185 (4.8) | 3852 (4.5) | 764 (3.2) | 486 (1.9) | 953 (2.5) | 2925 (4.0) | 3077 (4.1) | 9925 (5.0) | 48534 (6.5) |
| **Poverty level** |  |  |  |  |  |  |  |  |  |
| ≤100 | 100783 (79.0) | 67759 (79.2) | 17760 (74.2) | 18945 (73.2) | 28043 (73.6) | 57416 (79.2) | 58970 (78.3) | 153124 (76.6) | 511662 (68.6) |
| 101-150 | 7301 (5.7) | 4483 (5.2) | 1496 (6.2) | 1633 (6.3) | 2384 (6.3) | 3539 (4.9) | 4144 (5.5) | 11766 (5.9) | 52920 (7.1) |
| 151-200 | 2791 (2.2) | 1685 (2.0) | 564 (2.4) | 732 (2.8) | 897 (2.4) | 1254 (1.7) | 1477 (2.0) | 4822 (2.4) | 26311 (3.5) |
| >200 | 3478 (2.7) | 2350 (2.7) | 1051 (4.4) | 1342 (5.2) | 1750 (4.6) | 1914 (2.6) | 2181 (2.9) | 7263 (3.6) | 48386 (6.5) |
| Unknown | 13258 (10.4) | 9278 (10.8) | 3078 (12.9) | 3220 (12.4) | 5052 (13.3) | 8373 (11.5) | 8563 (11.4) | 22945 (11.5) | 106696 (14.3) |
| **Encounter type** |  |  |  |  |  |  |  |  |  |
| Ambulatory | 123670 (96.9) | 82126 (96.0) | 23181 (96.8) | 24816 (95.9) | 36458 (95.6) | 69476 (95.8) | 72290 (96.0) | 193044 (96.6) | 722744 (96.9) |
| Telehealth | 1257 (1.0) | 1084 (1.3) | 205 (0.9) | 417 (1.6) | 580 (1.5) | 906 (1.2) | 937 (1.2) | 2349 (1.2) | 11380 (1.5) |
| Unknown | 2684 (2.1) | 2345 (2.7) | 563 (2.4) | 639 (2.5) | 1088 (2.9) | 2114 (2.9) | 2108 (2.8) | 4527 (2.3) | 11851 (1.6) |
| **Health insurance** |  |  |  |  |  |  |  |  |  |
| Medicaid | 87323 (68.4) | 57845 (67.6) | 15525 (64.8) | 17238 (66.6) | 23709 (62.2) | 48430 (66.8) | 49227 (65.3) | 133126 (66.6) | 445445 (59.7) |
| Medicare | 11903 (9.3) | 7360 (8.6) | 3208 (13.4) | 2565 (9.9) | 4560 (12.0) | 7718 (10.6) | 7551 (10.0) | 20521 (10.3) | 80835 (10.8) |
| Private | 9713 (7.6) | 5960 (7.0) | 2230 (9.3) | 2899 (11.2) | 3999 (10.5) | 4385 (6.0) | 5892 (7.8) | 18100 (9.1) | 127963 (17.2) |
| Uninsured | 14294 (11.2) | 10870 (12.7) | 2157 (9.0) | 2329 (9.0) | 4485 (11.8) | 8997 (12.4) | 9489 (12.6) | 21272 (10.6) | 70495 (9.5) |
| Other Public | 1515 (1.2) | 1053 (1.2) | 238 (1.0) | 170 (0.7) | 242 (0.6) | 761 (1.0) | 942 (1.3) | 2122 (1.1) | 8072 (1.1) |
| No Information | 2863 (2.2) | 2467 (2.9) | 591 (2.5) | 671 (2.6) | 1131 (3.0) | 2205 (3.0) | 2234 (3.0) | 4779 (2.4) | 13165 (1.8) |
| **Facility type** |  |  |  |  |  |  |  |  |  |
| Ancillary | 29946 (23.5) | 19265 (22.5) | 4277 (17.9) | 4733 (18.3) | 7082 (18.6) | 16550 (22.8) | 17237 (22.9) | 43306 (21.7) | 121870 (16.3) |
| Medical | 94909 (74.4) | 63898 (74.7) | 19082 (79.7) | 20486 (79.2) | 29922 (78.5) | 53789 (74.2) | 55938 (74.3) | 151974 (76.0) | 612003 (82.0) |
| Specialty | 72 (0.1) | 47 (0.1) | 27 (0.1) | 14 (0.1) | 34 (0.1) | 43 (0.1) | 52 (0.1) | 113 (0.1) | 251 (0.0) |
| Unknown | 2684 (2.1) | 2345 (2.7) | 563 (2.4) | 639 (2.5) | 1088 (2.9) | 2114 (2.9) | 2108 (2.8) | 4527 (2.3) | 11851 (1.6) |

#### Transformation of potential predictors

Categorical variables were converted into binary formats using one-hot encoding to enhance the performance of ML models. This transformation simplified the dataset and reduced potential modeling complexities, making it more amenable to the ML algorithms used in our analysis [28–30]. After one-hot encoding, categories with very low prevalence were excluded due to near-zero variance, as such features provide minimal information for model training. Age was included as a normalized continuous variable without polynomial or binning transformations. The final feature set consisted of 13 predictor variables, which were used for all model training and evaluation.

#### Construction of datasets

Given that we were interested in predicting seven HRSN domains, plus the composite of overall HRSNs, we constructed eight datasets for each prediction. This was necessary because a significant proportion of the patient population were screened for only one HRSN domain. Thus, each HRSN domain dataset comprised of only those patients who had been screened for that domain.

### Development of ML Models

Model performance was evaluated using 10-fold stratified cross-validation on the full dataset, using SMOTE (Synthetic Minority Over-sampling Technique) [31] within the training portion of each fold to address class imbalance. Training was performed by one baseline statistical model, logistic regression, and three supervised ML algorithms: random forest (RF), Extreme Gradient Boosting (XGBoost), and Light Gradient-Boosting Machine (LightGBM). These algorithms were selected based on their demonstrated strengths in predictive modeling. Furthermore, they offer a balance of accuracy, efficiency, and interpretability, enabling robust and reliable prediction across diverse datasets [32–34]. Logistic regression was included as a baseline statistical model due to its widespread use and interpretability. L2 regularization was used in the model estimation to address potential multicollinearity from one-hot encoded variables. RF is an ensemble method that aggregates multiple decision trees. It is well-regarded for its robustness to overfitting, effective handling of high-dimensional data, and ability to estimate feature importance [35]. XGBoost is a gradient-boosting algorithm known for its superior predictive performance, computational efficiency, and ability to handle missing values, making it particularly suitable for complex datasets [36]. LightGBM is optimized for speed and scalability. It efficiently handles large-scale and high-dimensional data through histogram-based algorithms and direct support for categorical features [37].

Optimal hyperparameters for each model were identified using the 10-fold cross-validation process combined with a grid search on the training data. This process was applied within each cross-validation fold during model development. The primary evaluation metric for the trained models was the area under the receiver operating characteristic (AUROC) curve. AUROC provides a comprehensive measure of the model’s discriminatory performance by assessing the trade-off between sensitivity (true positive rate) and specificity (false positive rate) across different classification thresholds [38, 39]. It helps to evaluate how well the model distinguishes between positive and negative cases, independent of class prevalence [40]. Model performance was evaluated using conventional AUROC thresholds commonly applied in clinical prediction, with values near 0.50 indicating no discriminatory ability, 0.50-0.70 considered poor, 0.70-0.80 acceptable, 0.80-0.90 excellent, and >0.90 outstanding [38, 41]. For descriptive purposes, we refer to the upper end of the 0.5-0.7 range as modest, particularly when comparing relative improvements. Prior work has shown that even small absolute increases in AUROC on the order of 0.03-0.05 may correspond to meaningful improvements in discriminatory performance [39]. Beyond AUROC, we also measured model accuracy and recall. Accuracy provides an overall measure of correct predictions, while recall (sensitivity) specifically evaluates the model’s ability to identify patients with an unmet HRSN. Recall was included to ensure that the models were not biased toward predicting a lack of HRSNs for most cases, which could lead to high accuracy but poor identification of true positives. In our experience, models that do not account for this often achieve misleadingly high accuracy while remaining clinically unusable.

### Model Explainability

To enhance the interpretability of the ML models, we used SHAP (SHapley Additive exPlanations) [42, 43]. For SHAP analysis, the LightGBM model was trained using a 70% training and 30% testing split, and SHAP values were generated using the test set. SHAP elucidated the relative contribution of each predictor to the models’ predictions, providing a global understanding of feature importance and interactions. SHAP summary plots were generated to provide visual insights into the direction and magnitude of each predictor’s contribution to the prediction of each outcome.

## RESULTS

Of the 745,975 patients who were screened for at least one of the seven HRSNs, 26.8% had at least one unmet HRSN. Food insecurity (23.8%) was the most prevalent unmet HRSN as compared to the rest - utility needs (19.7%), housing instability (18.4%), social isolation (17.3%), transportation barriers (16.2%), poor housing quality (7.9%), and relationship safety (6.5%). The distribution of potential predictors by the overall and domain-specific HRSNs is reported in Table 1. The mean age of patients in the study was 37.4 years (SD = 19.4), with 62.6% identifying as female. White and Black patients comprised 57.3% and 23.1% of the study population, respectively. Overall, 68.6% of patients were at or below 100% of the FPL. Health insurance status included 59.7% with Medicaid, 10.8% with Medicare, and 9.5% who were uninsured. Detailed sociodemographic characteristics of the patients are presented in Table 1.

### Model Performance

The predictive performance (AUROC) and accuracy of the ML models are presented in Table 2. The ability of all three models to distinguish between those with any unmet HRSNs vs those without was modest [44], although LightGBM (64.5%) slightly outperformed logistic regression (61.4%), RF (63.7%), and XGBoost (60.3%). The performance advantage of LightGBM compared to the logistic regression baseline was statistically significant based on a paired Wilcoxon signed-rank test using 10-fold cross validation (*P*=.002). A similar pattern of model performance was observed for the domain-specific unmet HRSN models; LightGBM consistently outperformed logistic regression, RF, and XGBoost. The accuracies of the models were also moderate for the overall HRSN model – 54.9% (logistic regression), 58.7% (RF), 58.4% (XGBoost), and 59.5% (LightGBM). Similar moderate accuracy was observed for the prediction on domain-specific unmet HRSNs. The recall of all models, i.e. the probability of accurately identifying true cases of unmet HRSNs, was modest to acceptable overall and by each domain of unmet HRSNs. Additional performance metrics, including precision, are provided in the supplementary materials.

**Table 2.** Predictive performance of baseline and machine learning models for overall and domain-specific health-related social needs (HRSNs) among OCHIN network patients screened between January 1, 2016, and December 31, 2022. Four models (logistic regression, random forest, XGBoost, and LightGBM) were trained separately for each HRSN domain and for overall HRSN status, which was defined as having at least one unmet need in any of the seven domains. Model performance was evaluated using 10-fold cross-validation, and metrics reported include area under the receiver operating characteristic curve (AUROC), accuracy, and recall.

| HRSN Domain | Model | AUROC, % | Accuracy, % | Recall, % |
| --- | --- | --- | --- | --- |
| Overall | Logistic regression | 61.4 | 54.9 | 66.9 |
|  | Random forest | 63.7 | 58.7 | 63.0 |
|  | XGBoost | 60.3 | 58.4 | 64.6 |
|  | LightGBM | 64.5 | 59.5 | 62.7 |
| Food Insecurity | Logistic regression | 58.9 | 48.1 | 73.6 |
|  | Random forest | 61.3 | 54.7 | 65.3 |
|  | XGBoost | 58.7 | 53.2 | 69.9 |
|  | LightGBM | 62.3 | 54.8 | 67.3 |
| Housing Instability | Logistic regression | 61.6 | 52.1 | 70.3 |
|  | Random forest | 64.0 | 57.7 | 65.1 |
|  | XGBoost | 60.8 | 58.0 | 65.5 |
|  | LightGBM | 65.0 | 58.9 | 64.4 |
| Housing Quality | Logistic regression | 60.7 | 46.7 | 72.3 |
|  | Random forest | 59.8 | 53.5 | 62.8 |
|  | XGBoost | 59.1 | 50.2 | 69.6 |
|  | LightGBM | 61.9 | 51.6 | 67.9 |
| Relationship Safety | Logistic regression | 66.0 | 50.6 | 76.0 |
|  | Random forest | 65.7 | 56.9 | 68.8 |
|  | XGBoost | 63.0 | 52.4 | 75.1 |
|  | LightGBM | 67.5 | 56.5 | 71.0 |
| Social Isolation | Logistic regression | 61.3 | 55.6 | 64.2 |
|  | Random forest | 61.2 | 55.6 | 64.6 |
|  | XGBoost | 59.9 | 53.1 | 70.5 |
|  | LightGBM | 62.9 | 53.7 | 69.7 |
| Transportation | Logistic regression | 62.4 | 51.5 | 72.9 |
|  | Random forest | 64.0 | 58.3 | 64.3 |
|  | XGBoost | 61.2 | 57.5 | 67.0 |
|  | LightGBM | 65.0 | 59.3 | 64.5 |
| Utilities | Logistic regression | 60.5 | 50.9 | 68.4 |
|  | Random forest | 62.4 | 55.6 | 65.1 |
|  | XGBoost | 59.8 | 55.7 | 66.7 |
|  | LightGBM | 63.5 | 56.9 | 64.8 |

### Model Interpretation using SHAP

Given that LightGBM slightly outperformed the baseline logistic regression, as well as RF and XGBoost, we focused on interpreting the LightGBM models. The relative contribution of each model towards predicting the presence of any HRSN (overall) is illustrated with a SHAP summary plot as shown in Figure 1. The horizontal axis represents SHAP values, indicating the effect of each feature on the predicted outcome, with positive values increasing the likelihood of an HRSN and negative values decreasing it. Each point represents a SHAP value for a patient, showing how much a given feature contributed to increasing or decreasing the predicted value. Sample points are colored by the feature value (red = higher values or binary yes/1; blue = lower values or binary no/0). The vertical axis lists the features included in the model, sorted by importance.

**Figure 1.**
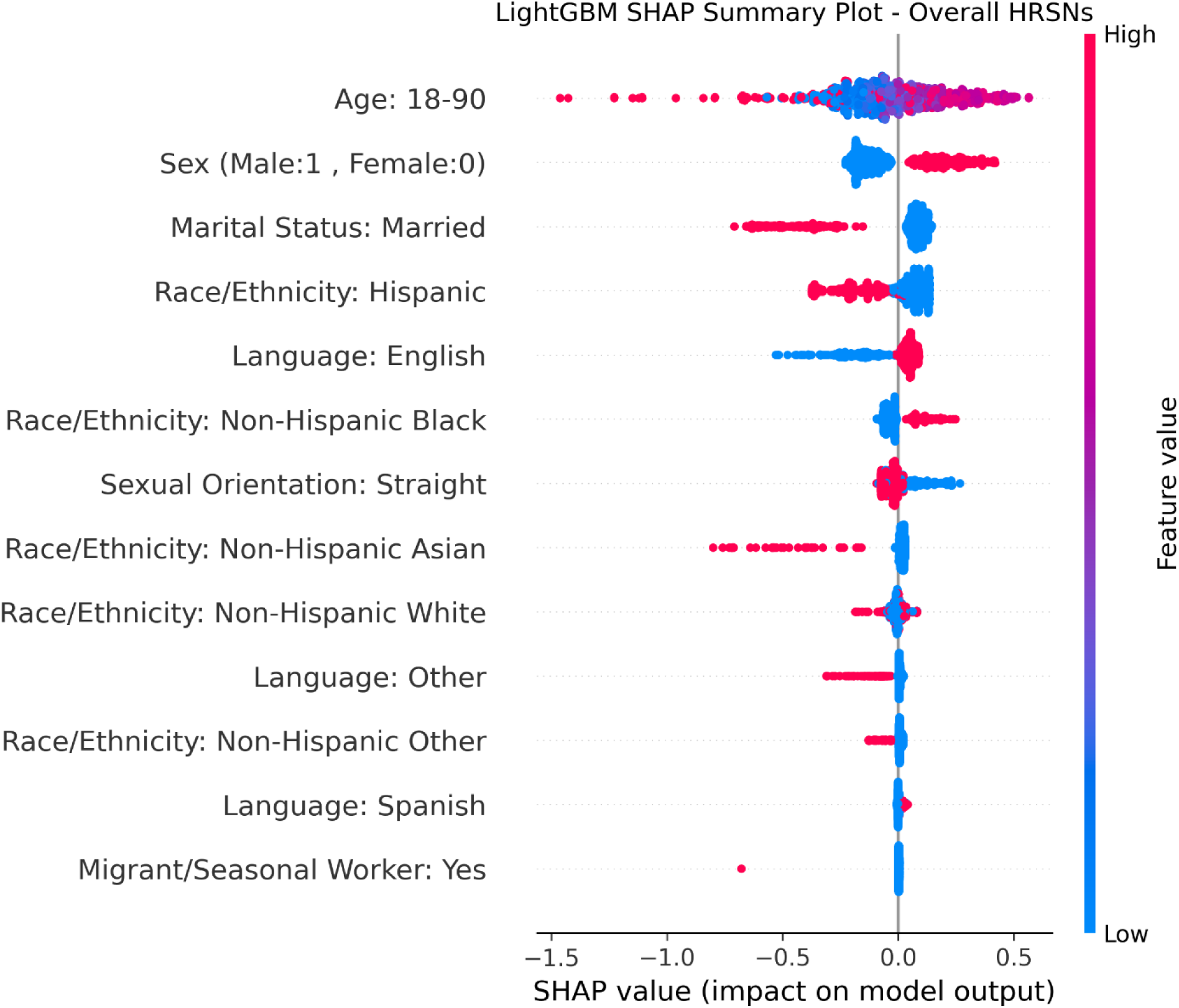
Exemplar SHAP summary plot highlighting feature contributions to overall HRSN prediction using the LightGBM model among OCHIN network patients screened between January 1, 2016, and December 31, 2022. Overall HRSN status was defined as having at least one unmet need in any of the seven domains: food insecurity, housing instability, poor housing quality, relationship safety, social isolation, transportation barriers, and utility needs. The horizontal axis represents SHAP values, where positive values indicate an increased likelihood of the outcome and negative values indicate a decreased likelihood. The vertical axis lists the features included in the model, sorted by importance. Samples are colored according to their feature value: red indicates higher values or a binary yes/1, and blue indicates lower values or a binary no/0. For the age feature, samples corresponding to younger and older individuals (blue and red) are predominantly associated with lower SHAP values, whereas samples corresponding to midlife (purple) show higher positive SHAP contributions.

The top-most features influencing the prediction of any HRSN were age, sex, marital status, race/ethnicity, and language. Specifically, SHAP values indicated that age contributed most strongly to model predictions among middle-aged patients relative to younger or older age groups. Other patterns included an increased likelihood for male, non-Hispanic Black, and English as a first language, and a decreased likelihood for married, Hispanic, non-Hispanic Asian, non-Hispanic Other, straight, or languages other than English as a first language. Similar patterns of feature importance were observed for each domain-specific model (*Supplementary Figures e1 – e7*). Readers should note that these figures are intended to demonstrate model interpretability and illustrate how predictions are generated, rather than to quantify social risk.

## DISCUSSION

### Principal Findings

Using data from patients who had been screened for at least one HRSN in the OCHIN EHR database, we sought to develop ML models for identifying patients with unmet HRSNs using a limited set of unmodifiable sociodemographic factors that are routinely collected in EHR databases. Overall, nearly a third of the patients who received care at any of the CHCs in OCHIN reported experiencing at least one unmet HRSN in the past month. The majority of these patients reported experiencing food insecurity in the past month. The ability of the resultant ML models (LightGBM) to identify correctly the presence of unmet HRSNs among patients was modest (AUROC range: 62% - 68%). Race/ethnicity, age, sex, marital status, and language were observed as the major influencers in the overall HRSN model. SHAP-derived feature importance rankings should be interpreted as hypothesis-generating, rather than as guides to resource allocation decisions. The modest predictive performance of these models suggests that, while these factors do contribute to identifying patients with unmet HRSNs, they are insufficient on their own to predict these needs fully. However, these findings define an upper bound on HRSN prediction performance using only non-modifiable demographic EHR data, serving as a baseline for future modeling efforts.

### Comparison to Prior Work

The predictive performance of our models, overall and by HRSN domains, were on par with similar HRSN prediction models previously described by Holcomb et al. [27] and Grant et al. [45], both of which reported peak AUROC scores of approximately 68%. While all three studies included demographic features, the Holcomb et al. study further included disease diagnoses, medical procedures, and insurance types, while the Grant et al. study included a broader set of EHR variables covering such things as diagnoses, behavior, insurance, neighborhood, and pharmacy, plus public census tract data. Furthermore, while our study included only individual-level features from EHR, the Holcomb et al. study included community-level SDOH (census-level needs for housing, food, transportation, utilities, and HRSNs) as features in their models. One possible reason our models achieved comparable performance, despite using fewer EHR features, may be that they included healthcare utilization features, such as disease diagnoses and medical procedures. Healthcare utilizations are more likely to be predicted by unmet HRSNs, rather than the other way round. Thus, we suggest this as a hypothesis worthy of examination in subsequent research.

### Future Directions

While the predictive abilities of our models were modest based on the inclusion of only a few non-modifiable sociodemographic features, it is plausible that we could potentially further improve the performance of our models by including features from the broader environments where patients live and work (i.e., contextual SDOH). Our ongoing efforts aim to explore the inclusion of hundreds of these contextual SDOH using environmental, built, and social exposomic variables, employing techniques that would help us overcome some of the limitations of feature selection in the current and prior studies [46, 47]. These future approaches should provide a more comprehensive assessment than models based on a limited set of non-modifiable features.

A critical next step will be external validation using data from an independent healthcare system, such as Heart of Ohio Family Health, which operates six Federally Qualified Health Centers based in Franklin County, Ohio. This would allow us to assess model transportability and recalibrate performance prior to broader deployment.

### Limitations

The primary limitation of this study is its lack of external validation, as the models were developed and tested on a single dataset. Their performance may therefore vary when applied to other populations or healthcare systems. Another limitation is the moderate predictive performances of our ML models. Given the overall modest results achieved, the false-positive rate may also be high. Early implementation without additional data enrichment could increase demands on resource-constrained clinics. As such, models would need to be refined and improved before adoption in real-world clinical settings. It should also be noted that our models relied solely on individual-level data from EHRs, which may constrain predictive accuracy. Finally, substantial missingness in variables such as migration/seasonal status may reflect site-level practices rather than true patient-specific absence. These variables demonstrated minimal contribution to model performance across domains, based on SHAP values, but would warrant further evaluation if future models exhibit stronger effects.

### Conclusions

Despite its limitations, this study also has several important strengths. First, our models used patient-level non-modifiable sociodemographic factors. Thus, our models leverage variables that are largely available in most EHR databases. At present, however, they are not ready for integration with clinical platforms. Second, our models were trained on one of the most diverse and most at-risk patient populations for unmet HRSNs across the nation. As such, this may support potential generalizability although external validation in independent healthcare settings is required before we can make such claims. Third, and unlike prior studies, we used SHAP to understand and interpret the potential drivers of unmet HRSNs. This is central to enhancing the interpretability and transparency of the models that can then help empower healthcare providers to understand and trust model predictions.

Overall, this study establishes a benchmark for the predictive performance achievable when utilizing only commonly available, non-modifiable sociodemographic data in EHR. Despite the promise of this approach, the predictive performance of the models we have employed must be improved before they are likely to see widespread adoption in real-world clinical settings.

## Funding Statement

This project (1OT2OD032581-02-394) is funded by the National Institutes of Health (NIH) through the Artificial Intelligence/Machine Learning Consortium to Advance Health Equity and Researcher Diversity (AIM-AHEAD) program.

## Acknowledgments

The research reported in this work was powered by PCORnet®. PCORnet has been developed with funding from the Patient-Centered Outcomes Research Institute® (PCORI®) and conducted with the Accelerating Data Value Across a National Community Health Center Network (ADVANCE) Clinical Research Network (CRN). ADVANCE is a Clinical Research Network in PCORnet® led by OCHIN in partnership with Health Choice Network, Fenway Health, University of Washington, and Oregon Health & Science University. ADVANCE’s participation in PCORnet® is funded through the PCORI Award RI-OCHIN-01-MC.

## Data Availability

The data that support the findings of this study are available from OCHIN, but restrictions apply to the availability of these data, which were used under license for the current study, and so are not publicly available. Data are, however, available from the authors upon reasonable request and with permission of OCHIN.

## Authors’ Contributions

Conceptualization: MD; Data curation: TA, YC; Formal analysis: LD, MI, MD; Funding acquisition: MD; Investigation: LD; Methodology: LD, MI; Project administration: MD; Resources: MAL, MD; Software: LD; Supervision: MAL, MD; Validation: LD; Visualization: LD; Writing – original draft: LD, MI, MD; Writing – review & editing: TA, YC, AM, AD, BM, MAL

## Additional Information

The authors declare no competing interests.

## Notes

### Competing Interest Statement

The authors have declared no competing interest.

### Author Declarations

This study was approved by The Ohio State University Institutional Review Board (IRB). The requirement for patient informed consent was waived by The Ohio State University IRB because the data were de-identified by OCHIN and posed minimal risk of re-identification. This study was reported in accordance with the Transparent Reporting of a Multivariable Prediction Model for Individual Prognosis or Diagnosis (TRIPOD) statement. All methods were performed in accordance with the relevant guidelines and regulations.

