## Supplemental Figures and Table for "Leveraging Machine Learning Approaches to Identify Health-Related Social Needs Screening from Electronic Health Records"

### Supplementary Figures

**Figure e1:** SHAP summary plot for housing instability

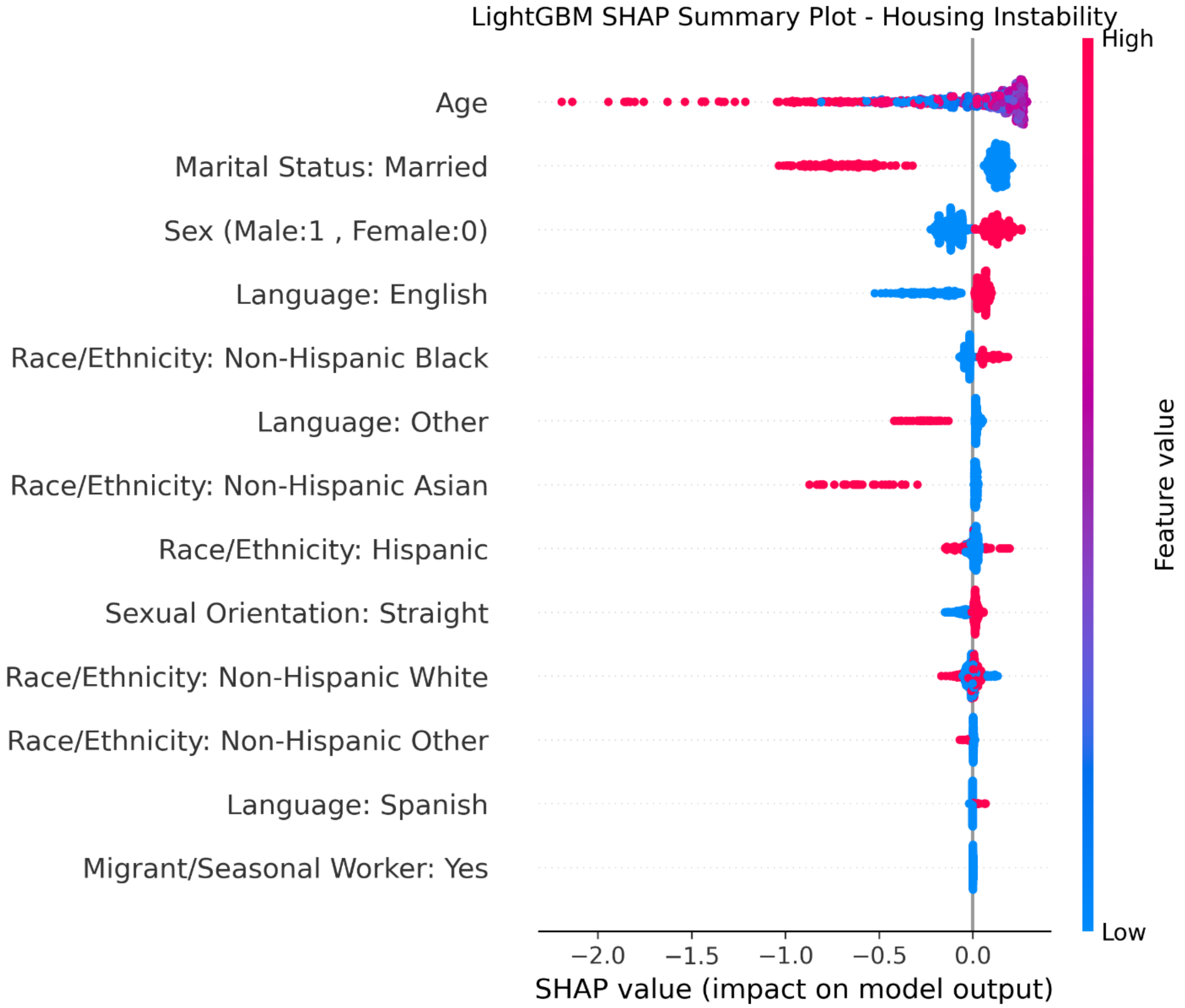

**Figure e2:** SHAP summary plot for housing quality

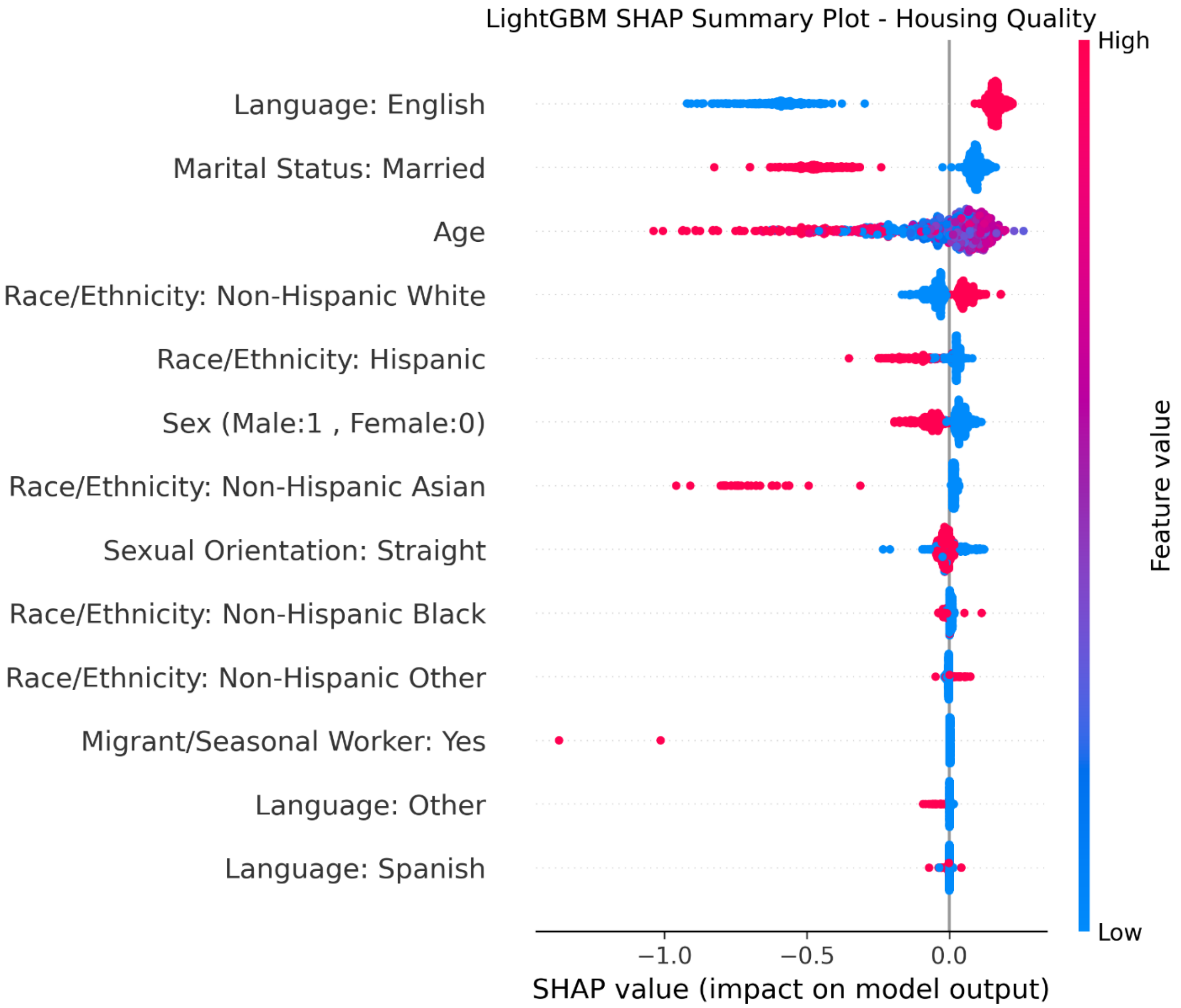

**Figure e3**: SHAP summary plot for relationship safety

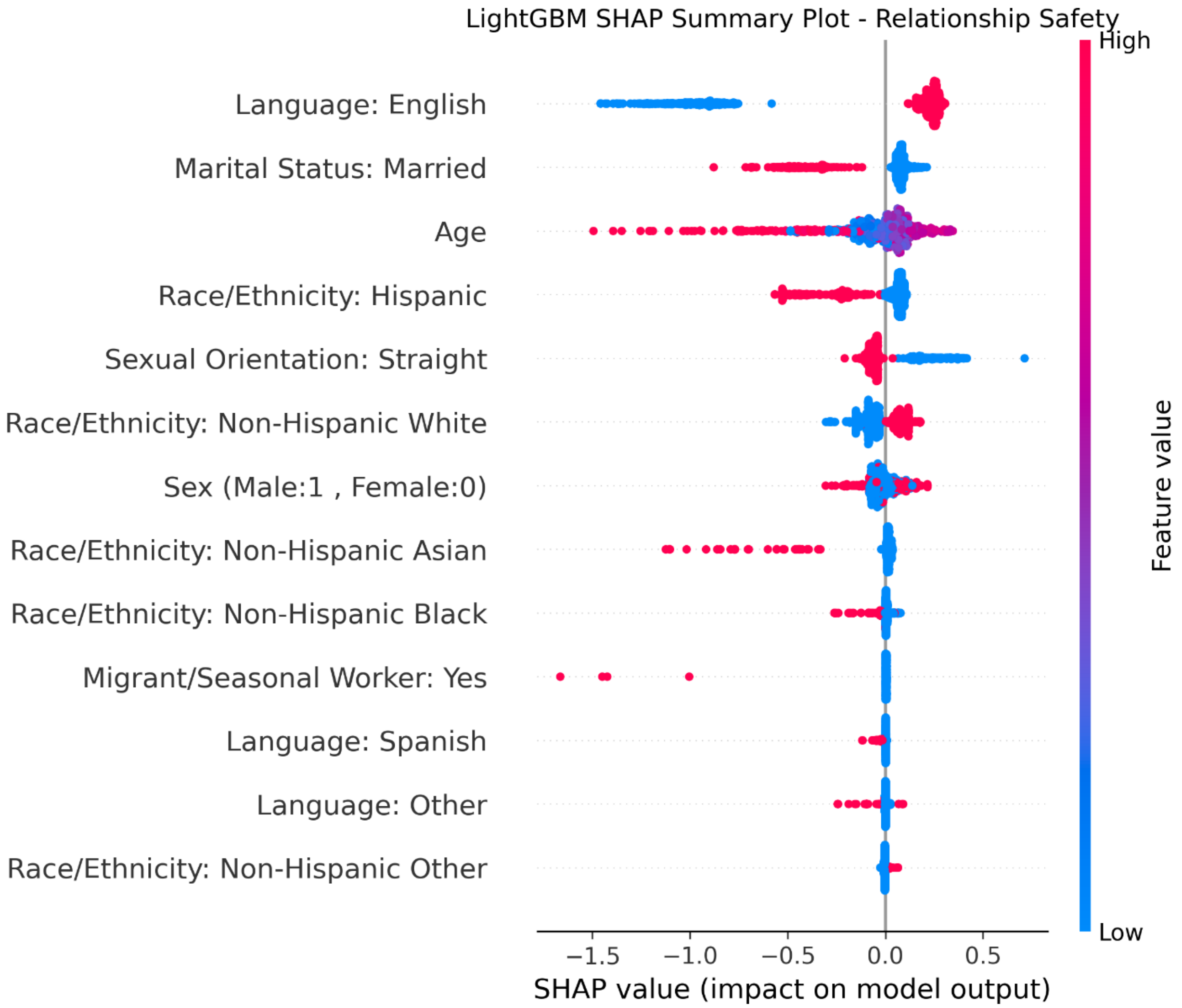

**Figure e4**: SHAP summary plot for social isolation

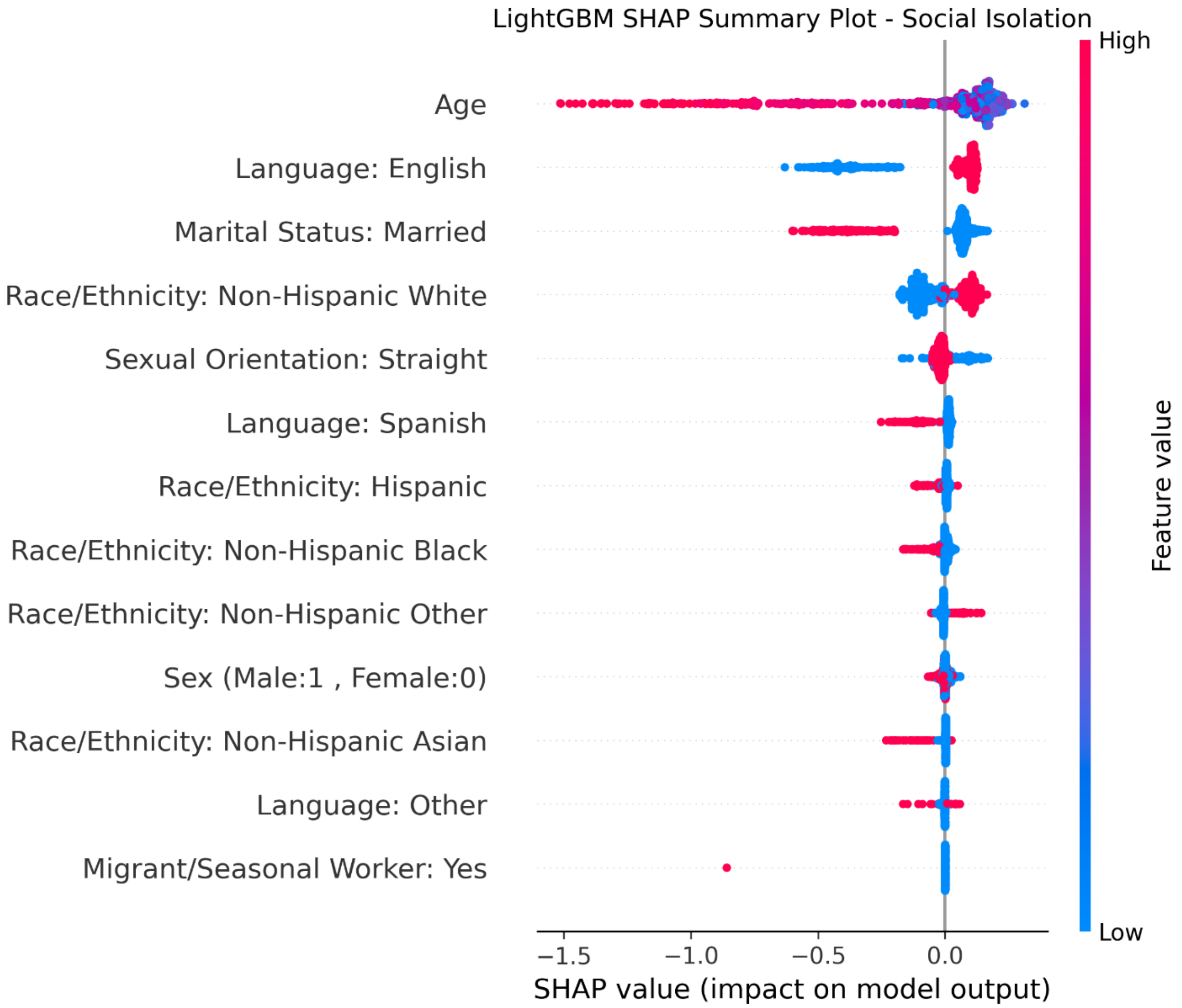

**Figure e5:** SHAP summary plot for transportation

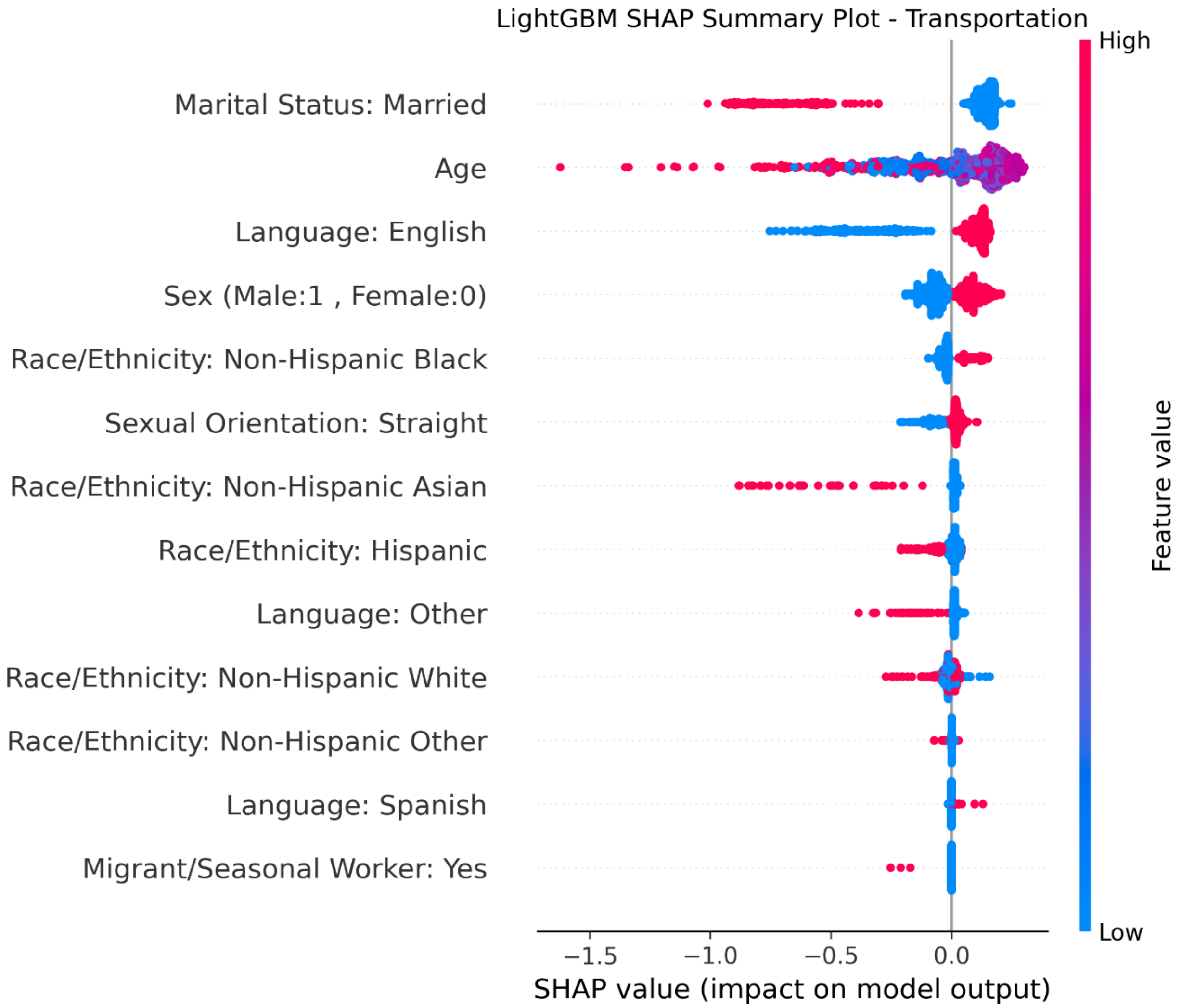

**Figure e6**: SHAP summary plot for utilities

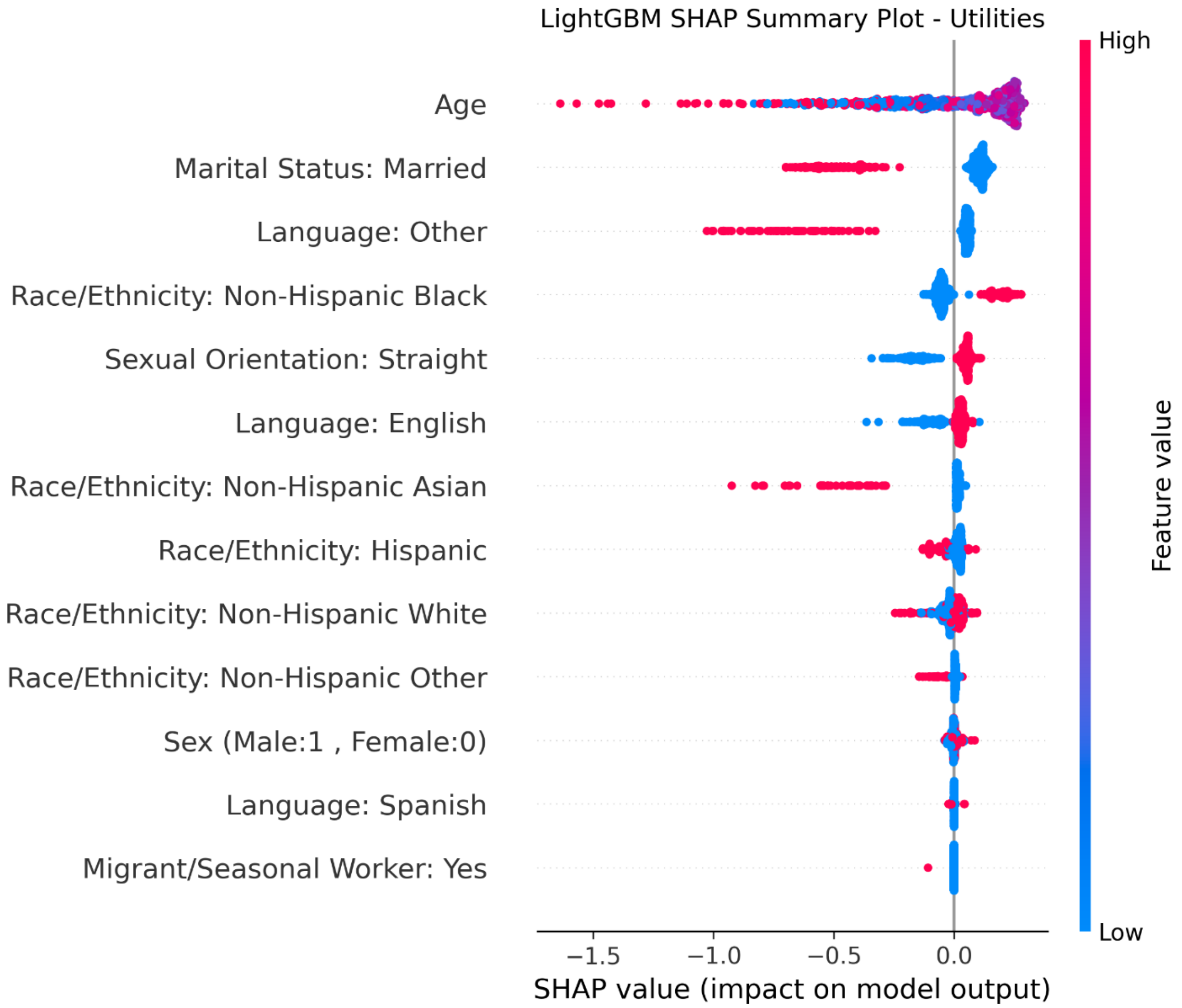

**Figure e7**: SHAP summary plot for food insecurity

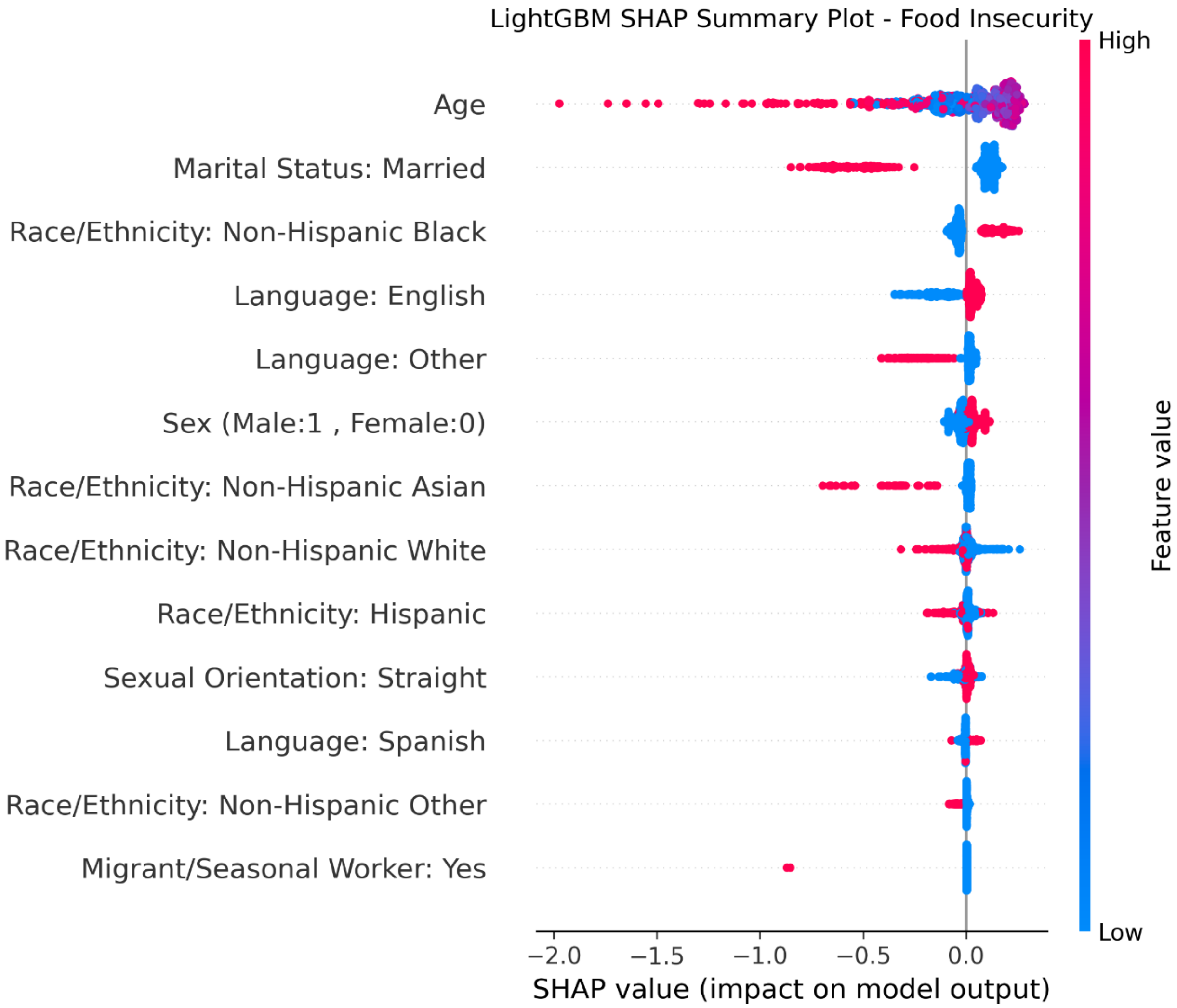

### Supplementary Table

**Table e1:** Complete performance metrics for all models and HRSN domains, including AUROC, accuracy, precision, recall, and F1 score.

| HRSN Domain | Model | AUROC, % (95% CI) | Accuracy, % | Precision, % | Recall, % | F1, % |
|---|---|---|---|---|---|---|
| **Overall** | Logistic regression | 61.37 (61.10, 61.67) | 54.90 | 34.03 | 66.86 | 45.11 |
| | Random forest | 63.67 (63.40, 63.88) | 58.69 | 35.99 | 62.99 | 45.81 |
| | XGBoost | 60.33 (60.13, 60.64) | 58.42 | 36.05 | 64.62 | 46.28 |
| | LightGBM | 64.53 (64.32, 64.74) | 59.45 | 36.52 | 62.72 | 46.16 |
| **Food Insecurity** | Logistic regression | 58.92 (58.44, 59.44) | 48.08 | 28.92 | 73.57 | 41.52 |
| | Random forest | 61.29 (60.84, 61.69) | 54.72 | 30.90 | 65.33 | 41.95 |
| | XGBoost | 58.73 (58.36, 59.15) | 53.15 | 30.82 | 69.91 | 42.78 |
| | LightGBM | 62.33 (61.94, 62.81) | 54.76 | 31.27 | 67.25 | 42.69 |
| **Housing Instability** | Logistic regression | 61.64 (61.30, 62.36) | 52.05 | 24.94 | 70.25 | 36.82 |
| | Random forest | 64.02 (63.45, 64.84) | 57.66 | 26.77 | 65.07 | 37.93 |
| | XGBoost | 60.83 (60.39, 61.72) | 58.02 | 27.05 | 65.49 | 38.29 |
| | LightGBM | 65.03 (64.27, 65.95) | 58.89 | 27.35 | 64.44 | 38.40 |
| **Housing Quality** | Logistic regression | 60.71 (59.76, 61.31) | 46.69 | 10.09 | 72.30 | 17.71 |
| | Random forest | 59.84 (58.64, 60.37) | 53.48 | 10.26 | 62.79 | 17.64 |
| | XGBoost | 59.07 (57.98, 59.89) | 50.21 | 10.44 | 69.59 | 18.15 |
| | LightGBM | 61.86 (60.99, 62.85) | 51.58 | 10.51 | 67.90 | 18.20 |
| **Relationship Safety** | Logistic regression | 65.95 (65.37, 66.68) | 50.63 | 9.28 | 75.98 | 16.54 |
| | Random forest | 65.72 (65.14, 66.37) | 56.86 | 9.73 | 68.83 | 17.04 |
| | XGBoost | 62.98 (62.55, 63.61) | 52.40 | 9.51 | 75.13 | 16.89 |
| | LightGBM | 67.51 (66.94, 68.28) | 56.47 | 9.89 | 71.01 | 17.36 |
| **Social Isolation** | Logistic regression | 61.31 (60.67, 61.93) | 55.55 | 22.85 | 64.24 | 33.71 |
| | Random forest | 61.21 (60.78, 61.74) | 55.59 | 22.93 | 64.57 | 33.84 |
| | XGBoost | 59.93 (59.42, 60.58) | 53.08 | 22.91 | 70.51 | 34.58 |
| | LightGBM | 62.86 (62.31, 63.38) | 53.70 | 23.03 | 69.67 | 34.61 |
| **Transportation** | Logistic regression | 62.41 (61.95, 62.71) | 51.54 | 22.59 | 72.92 | 34.50 |
| | Random forest | 64.03 (63.65, 64.30) | 58.25 | 24.07 | 64.33 | 35.03 |
| | XGBoost | 61.23 (60.92, 61.42) | 57.50 | 24.19 | 66.95 | 35.54 |
| | LightGBM | 65.03 (64.65, 65.45) | 59.34 | 24.67 | 64.45 | 35.68 |
| **Utilities** | Logistic regression | 60.50 (60.15, 60.80) | 50.88 | 25.05 | 68.44 | 36.68 |
| | Random forest | 62.42 (61.71, 62.82) | 55.62 | 26.72 | 65.14 | 37.89 |
| | XGBoost | 59.77 (59.49, 60.22) | 55.72 | 27.07 | 66.71 | 38.51 |
| | LightGBM | 63.53 (63.10, 64.01) | 56.93 | 27.37 | 64.82 | 38.49 |
